# Validation of the PETS-stroke-A patient reported measure (PRM) of treatment burden after stroke

**DOI:** 10.1101/2024.11.21.24317754

**Authors:** Martin Taylor-Rowan, David T Eton, Hamish J. McLeod, Jala Rizeq, Lisa Kidd, Grace Currie, Terry J Quinn, Frances S Mair, Katie I Gallacher

## Abstract

**Introduction:** Treatment burden is the workload of healthcare for people with long-term conditions and the impact on wellbeing. A validated measure of treatment burden for use as an outcome measure in stroke trials is needed. We adapted a patient-reported measure (PRM) of treatment burden in multimorbidity, PETS (Patient Experience with Treatment and Self-Management version 2.0), to create a stroke-specific measure, PETS-stroke, and examined its psychometric properties.

**Patients and Methods:** We recruited stroke and transient ischaemic attack (TIA) survivors between Feb 2022-June 2023 from 10 hospitals in the UK and through the Scottish Health Research Register (SHARE). Participants completed the PETS-stroke questionnaire along with 3 other PRM’s (Stroke Southampton Self-Management Questionnaire, The Satisfaction with Stroke Care Measure, The Shortened Stroke Impact Scale). We performed confirmatory factor analysis to test the factor structure of the PETS-stroke. We assessed Spearman’s rank correlations between PETS-stroke and other PRMs to determine convergent validity. Intra-class coefficient was performed to assess test-retest reliability. Proportions of missing data along with feedback from qualitative interviews were used to determine feasibility. T-tests were conducted to examine variations in PETS-stroke scores based on multimorbidity and socioeconomic factors.

**Results:** Three-hundred-eighty-one participants were included. The best fit was achieved with a 9-factor structure and internal consistency was good (Omega values 0.729 to 0.921). The factor loadings for the individual indicator items across eight of the nine domains were moderate to strong. All domains of PETS-stroke showed moderate to strong correlations with at least one other PRM. Test-retest reliability was good for all domains (ICC>0.7). Qualitative feedback on feasibility was positive and missing data was within acceptable limits for 7 domains. PETS-stroke scores significantly differed based on multimorbidity in 3 domains and in 8 domains based on socioeconomic status.

**Discussion:** Psychometric performance suggests PETS-stroke is a highly promising measure of treatment burden after stroke.

## Introduction

The term *treatment burden* describes the healthcare workload of living with long-term conditions and the impact of this work on wellbeing. Treatments can become burdensome when there are too many, or if they are challenging to implement in everyday life. People with stroke describe the healthcare workload as draining of time, energy and finances.^1^ Additionally, stroke survivors may have physical, cognitive, or emotional impairments that can impede self-management.^2^ There has been a recent interest in understanding treatment burden and developing methods of measurement, to aid identification of high-risk groups and assist the testing of interventions.^3^ Understanding these issues in a sufficiently nuanced way will require the creation of condition-specific measures that have been developed and tested in line with best practice principles.^4^

Treatment burden is subjective; therefore, patient-reported methods are suitable for measurement. The Patient Experience with Treatment and Self-Management (PETS) 2.0^5^ is a commonly used patient-reported measure (PRM) of treatment burden; however it was designed for use in a general healthcare population and does not capture key aspects of the stroke treatment burden profile such as rehabilitation strategies for limb weakness or speech difficulties. Despite a distinct care pathway, no comprehensive PRMs of treatment burden in stroke currently exist.^6^ Consequently, we are unable to fully appreciate the experience of treatment burden amongst stroke survivors and the impact this may have upon their health outcomes.

To address this issue, we created the PETS-stroke, a highly modified version of the PETS 2.0, to measure treatment burden after stroke. The content development of PETS-stroke has been described previously.^7^ Here, we describe the evaluation of the psychometric properties of the tool. **Figure 1** presents the full course of PETS-stroke development highlighting the aspects reported here. Establishing the psychometric properties of PETS-stroke will determine whether the tool is ready to be used to measure treatment burden in stroke populations.

### Aims and objectives

Through patient surveys and interviews, our primary aims were:

1. To determine the factor structure of PETS-stroke (survey).
2. To assess the convergent validity of PETS-stroke (survey).
3. To assess the reliability of PETS-stroke (survey).
4. To assess the feasibility of PETS-stroke (survey and interviews).

Our secondary aims were to:

1. Establish known-groups validity of PETS-stroke, comparing PETS-stroke test scores with measures of socioeconomic status and multimorbidity (survey).
2. Establish user perspectives on potential alterations to future iterations of the measure (interviews).

**Figure 1.**
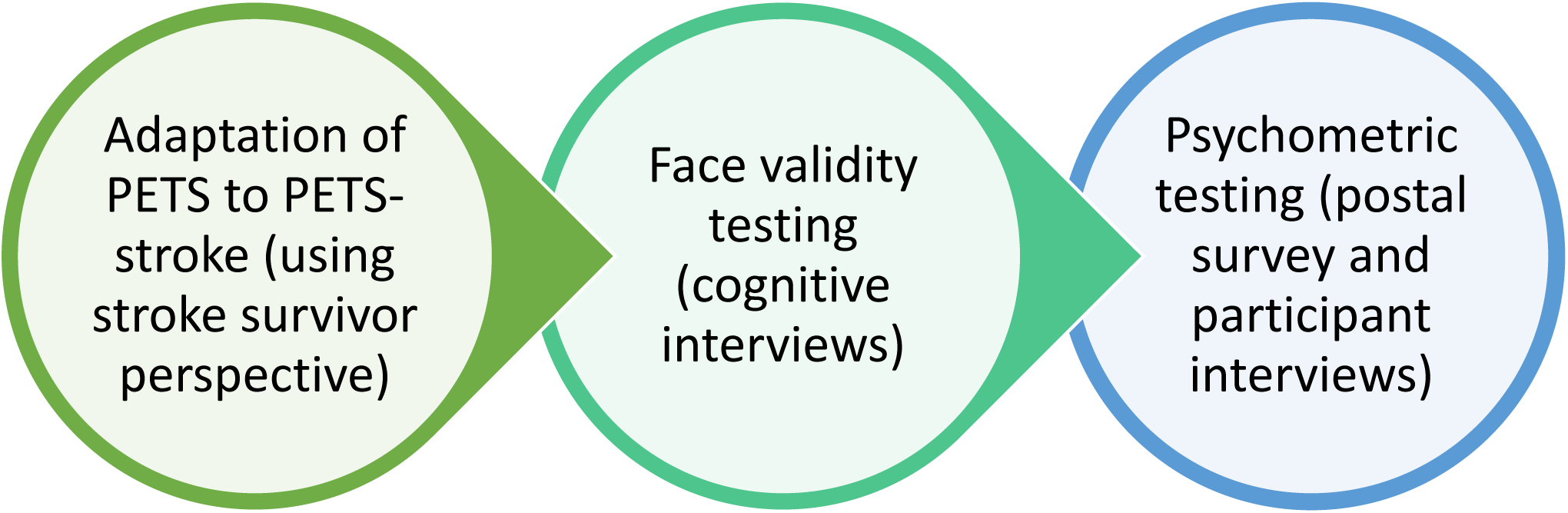
PETS-stroke development stages. PETS = Patient Experience with Treatment and Self-Management Measure

## Patients and Methods

The project was funded by the Chief Scientist Office (CSO) Scotland (HIPS/21/13). Ethical approval was provided by London and Surrey Borders NHS Research Ethics Committee (20/LO/0871). Guidance on PRM development published by the International Society for Quality of Life Research (ISOQOL)^8^ and COnsensus-based Standards for the selection of health Measurement INstruments (COSMIN)^9^ informed the methodology. Details of the methodology can be seen in the study protocol.^10^ A summary is given below.

### Survey methods

#### Recruitment

We recruited stroke and transient ischaemic attack (TIA) survivors between Feb 2022-June 2023 from 9 participating hospitals in Scotland, 1 in Wales, and through the Scottish Health Research Register (SHARE).^11^ Participants were eligible if they fulfilled the following criteria: community dwelling (including retirement housing or sheltered housing); stroke diagnosis (including ischaemic, haemorrhagic, any severity and TIA); over one month since hospital discharge, but less than 1 year post-stroke at time of completing the survey (in order to capture treatment burdens encountered at home rather than in the hospital, those not admitted to hospital were also included); able to read English; and able to complete the paper survey or ask someone to do as a proxy. We used a sampling frame to promote diversity in participant characteristics (age, sex, ethnicity, socioeconomic status, stroke sequalae, time since stroke).

#### Data collection

Stroke survivors were asked to complete survey packs containing the PETS-stroke measure and three additional PRMs: The Stroke Southampton Self-Management Questionnaire, measuring readiness to self-manage;^12^ The Satisfaction with Stroke Care Measure, measuring healthcare satisfaction;^13^ and The Shortened Stroke Impact Scale, measuring illness burden.^14^ In addition, self-reported demographic data were requested—details can be seen in the study protocol. ^15^

To determine test-retest reliability, the PETS-stroke was sent out a second time 2 weeks after completion of the first questionnaire and returned by the participant as soon as they were able.

#### Sample Size

Details of our sample size calculation are in our study protocol. In brief, our sample size calculation indicated that we required a total of 340 participants to give a ratio of approximately 10 participants per item and provide a comparable sample to the Confirmatory Factor Analysis (CFA) study of the PETS 2.0.^16^

A sample size calculation for follow-up indicated we would require 218 participants to provide questionnaires at both baseline and follow-up.

### Missing data

Pre-established scoring rules were followed for respective questionnaires (see **Supplementary materials 1**). For PETS-stroke similar rules were created based on the scoring system of the PETS 2.0 survey.^5^ Each domain is scored separately, and raw scores are standardised to generate scores ranging from 0-100 for each domain. Higher scores indicate greater treatment burden.

‘Not applicable’ responses were treated as missing values. PETS-stroke domains could be scored and included in analysis, despite missing data, provided >50% of items were answered within the domain. Otherwise, missing/not applicable demographic and questionnaire data were excluded from analyses.

#### Data analysis

All analyses and assumptions are described in a detailed *a priori* publicly available Statistical Analysis Plan (https://osf.io/6p2kt/).

#### Examination of sample characteristics

Sociodemographic characteristics of participants were reported descriptively.

#### Confirmatory Factor Analysis Model Specification, factor loadings and internal consistency reliability

The PETS-Stroke differed from the PETS-2.0; the scale was shortened to make it more tolerable for the target population and the item content and response scales were adapted based on qualitative data obtained from stroke survivors.^6^ The resulting 34-item measure shared some items and underlying theoretical constructs with the original scale. In order to pre-specify the factor structure of the PETS-Stroke, three subject matter experts (two general practitioners and one stroke specialist) independently reviewed the scale items and proposed sub-scale composition. Disagreements were then discussed and if needed the chief investigator of the study had the casting vote where consensus could not be reached (this only occurred on one occasion). Nine subscales were identified based on 33 items from the PETS-Stroke, with one remaining item assessing Mental Fatigue set aside as a single indicator item. The specification of this potential new structure was completed prior to estimating the models.

Based on the theoretical model of treatment burden and previous work on the PETS-2.0^16^ scale structure, CFA was used to test competing underlying models to assess the dimensionality of the new PETS-Stroke and identify the best fitting structure that would account for its revised item composition. Four models were estimated: 1) a one factor “treatment burden” model; 2) a bifactor (hierarchical) model with an overall treatment burden factor that loads onto all items and three orthogonal specific measurement factors representing workload, impact, and general burden (underlying items not captured by workload or impact); 3) a nine-factor model of the newly proposed structure; 4) a higher order model with one second order factor of treatment burden underlying the nine first order factors.

As the observed variables are measured on ordinal scales, analyses were conducted on polychoric correlation matrices and robust weighted least squares with adjustment for mean (WLSM) estimator was used. Model fit evaluation used multiple indices (model χ^2^, SRMR<0.08, RMSEA<0.06, TLI/CFI≥0.95) in line with best practice guidance.^17^ Internal consistency was assessed with McDonald’s omega based on realistic assumptions about the covariance between scale items and factors.^18–20^ Analyses were carried out in RStudio (version 2024.04.1+748) using the packages Lavaan (v 0.6-16) and semTools (v 0.5-6).

#### Evaluation of psychometric properties

To examine convergent validity, the PETS-stroke was compared to the 3 other PRMs using Spearman’s rank correlations, hypothesising that higher treatment burden would significantly correlate with lower readiness to self-manage, lower satisfaction with stroke services and increased burden of illness. For domains ‘keeping healthy’ and ‘living at home with stroke’, variation in the number of item responses within each domain skewed missing data imputation. To investigate the impact of this on results, a sensitivity analysis was performed restricting these domains to participants with full data only.

Test-retest reliability was assessed by calculating the intra-class coefficient (ICC) of baseline and follow-up assessments.

T-tests were performed to establish known-groups validity, comparing PETS-stroke test scores with socioeconomic status and multimorbidity. Socioeconomic status was measured using the Scottish Index of Multiple Deprivation (SIMD) and Welsh Index of Multiple Deprivation (WIMD) with scores in lowest and highest quintiles compared. Co-morbidities were self-reported, with a general practitioner interpreting results and generating a count of morbidities for each participant. Multimorbidity was defined as having ≥3 co-morbidities, with scores for this group compared to those with stroke only. We hypothesised that participants from more deprived socioeconomic areas and participants with multimorbidity would have significantly higher treatment burden. A sensitivity analysis was performed using Mann-Whitney U tests to examine known groups analysis to determine any impact of non-normal distribution.

#### Feasibility Testing

Scores were examined at domain level descriptively using range, means and standard deviations. Percentage of missing items and proportion of returned surveys provided information on acceptability. A total of ≥15% of participants being scored at the lowest or highest possible score for a domain was indicative of a possible floor/ceiling effect.

Semi-structured interviews were conducted by telephone with a subset of participants to enquire about usability of the survey—interview schedule is shown in **Supplementary materials 2**.^21^ Sampling was purposive, aiming for diversity in age, stroke severity, ethnicity and sex to include a broad range of viewpoints. Interviews were audio recorded and transcribed verbatim. Data was analysed using a codebook thematic analysis approach to look for key themes or topics arising,^22^ performed using NVivo software version 14.^23^

#### User perspectives on potential changes for future iterations

Authors discussed any possible alterations to PETS-stroke guided by the results of psychometric testing. Further interviews were conducted to gauge opinions on these potential changes. The same sampling, qualitative methodology, and analytical approach, described above, was employed.

## Results

### Sample characteristics

In total 396 participants were recruited. Fifteen were subsequently excluded due to incomplete consent forms, leaving 381 participants available for analysis. Participant demographics are shown in **Table 1**. Demographics were reasonably consistent with Scottish Stroke Care Audit statistics,^24^ with the study sample being slightly younger, more affluent, and more able-bodied than the national stroke average.

**Table 1.** Baseline demographics.

| Demographic | Number of valid cases (total=381) | Study descriptive data | National data- at first stroke assessment (n=11,257) |
| --- | --- | --- | --- |
| Age (Mean; SD) | 371 | 68.2 (11.24) | 73 |
| Gender (Female %*) | 378 | 43.3% | 47% |
| Ethnicity (%*) | 379 | White: 98.4%<br>Asian: 0.8%<br>Black: 0.3%<br>Other ethnic group: 0.3% | Not available |
| Education (%*) | 370 | None: 17.3%<br>School qualifications: 35.6%<br>Higher education: 46.8% | Not available |
| Number of co-morbidities (Mean; SD) | 379 | 2.2 (1.9) | Not available |
| Medication count (Mean; SD) | 375 | 5.8 (3.1) | Not available |
| Deprivation quintile (%*) | 372 | 1<sup>st</sup> (Most deprived): 17.5%<br>2<sup>nd</sup>: 18.8%<br>3<sup>rd</sup>: 17.2%<br>4<sup>th</sup>: 22.8%<br>5<sup>th</sup> (Least deprived): 23.1% | 1<sup>st</sup>(Most deprived): 24%<br>2<sup>nd</sup>: 22%<br>3<sup>rd</sup>: 20%<br>4<sup>th</sup>: 18%<br>5<sup>th</sup>(Least deprived): 16% |
| Speech difficulties (%*) | 380 | 20.3% | 29% |
| Difficulty using arms (%*) | 380 | 26.3% | 39% |
| Difficulty walking (%*) | 380 | 41.3% | 55% |
| Wheelchair use (%*) | 380 | 1.6% | Not available |
| Live alone (%*) | 379 | 26.3% | Not available |
| Have support with chores if needed (%*) | 374 | 92.2% | Not available |
| Participating in other research or trial (%) | 376 | 18.1% | Not available |

\*Percentages are rounded so ≠ 100%

### Factor analysis

#### Confirmatory Factor Analysis, factor loadings and internal consistency reliability

The CFA fit results including all fit indices are presented in **Table 2**. All models converged normally. A description of models 1,2 and 4 can be found in **Supplementary materials 3**.

**Table 2.** PETS-Stroke CFA model fit results (N= 378)

| *Models* | $\chi^2$ | df | CFI | TLI | RMSEA | 90%<br>CI | SRMR |
| --- | --- | --- | --- | --- | --- | --- | --- |
| Model 1: Single Factor Model | 6708.0 | 495 | 0.891 | 0.884 | 0.182 | 0.178-0.187 | 0.133 |
| Model 2: Bifactor Model | 2752.7 | 462 | 0.96 | 0.954 | 0.115 | 0.11-0.12 | 0.095 |
| **Model 3: Nine Factor Model** | **1710.0** | **459** | **0.978** | **0.975** | **0.085** | **0.079-**<br>**0.091** | **0.085** |
| Model 4: Higher Order Nine<br>Factor Model | 2018.2 | 486 | 0.973 | 0.971 | 0.091 | 0.086-0.097 | 0.092 |

*Notes:* All $\chi^2$ results are statistically significant ( $p < .001$ ); $\chi^2$ = chi-square test; df = degrees of freedom; CFI = Comparative Fit Index ( $> 0.90$ = adequate fit, $> 0.95$ = excellent fit); TLI = Tucker-Lewis Index ( $> 0.90$ = adequate fit, $> 0.95$ = excellent fit); RMSEA = Root Mean Square Error of Approximation ( $< 0.08$ = adequate fit, $< 0.06$ = excellent fit); SRMR = Standardised Root Mean Square Residual ( $< 0.1$ = acceptable; $< 0.08$ = good). Best-fitting model in **bold**

Model 3 was based on the assessment of our subject matter experts and this produced the best fitting model with all good or excellent values for all fit indices. **Table 3** provides the subscale names and items and we have presented more details on the model properties in the following sections.

The ω values for the nine domain scores ranged from 0.729 to 0.921 – all well above the thresholds required for acceptable (0.65) to strong (0.80) internal consistency.^19, 20^ The factor loadings for the individual indicator items across eight of the nine domains were moderate to strong. Two items in the “Living at Home with Stroke” domain showed factor loadings lower than .40.

**Table 3.**
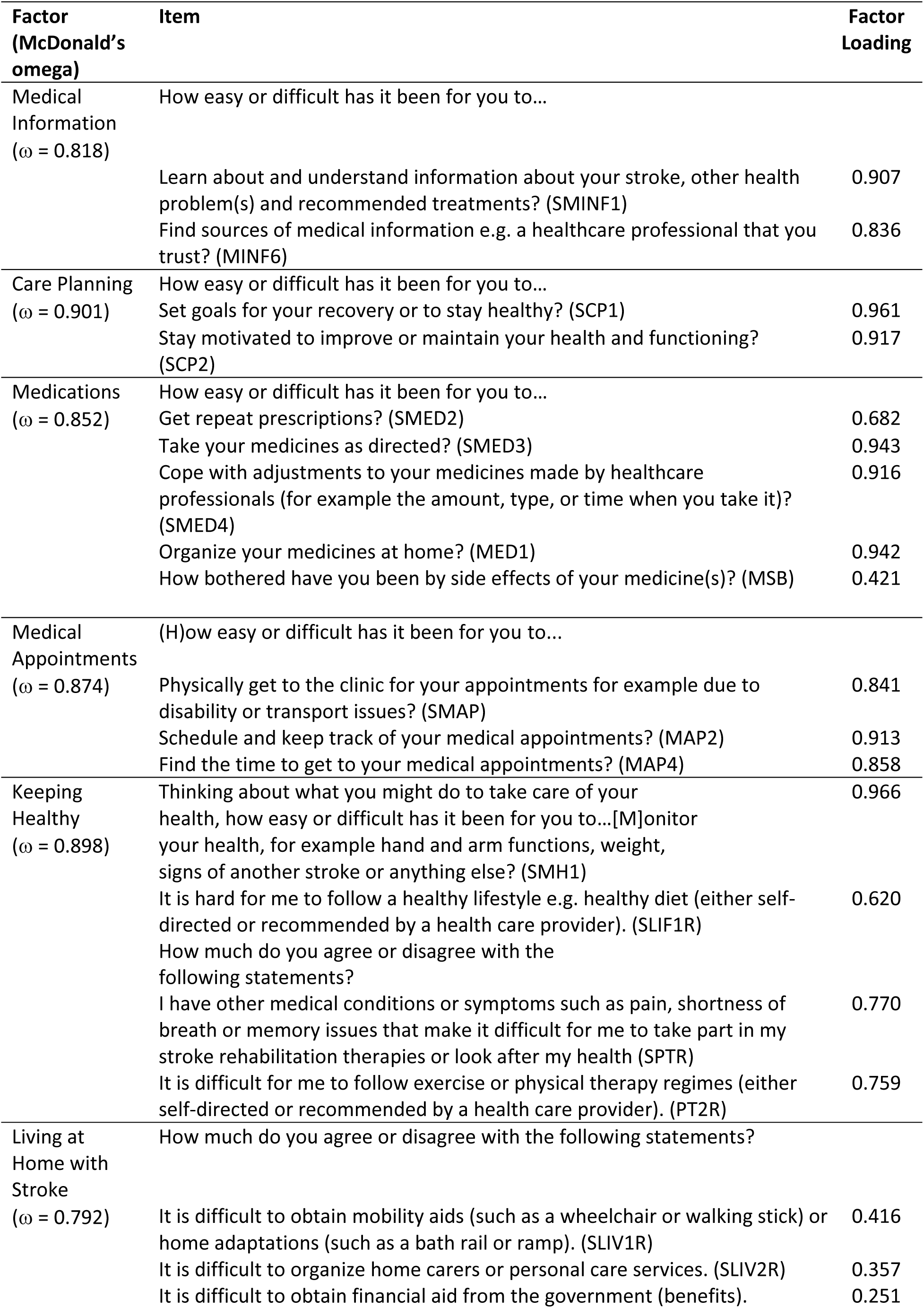
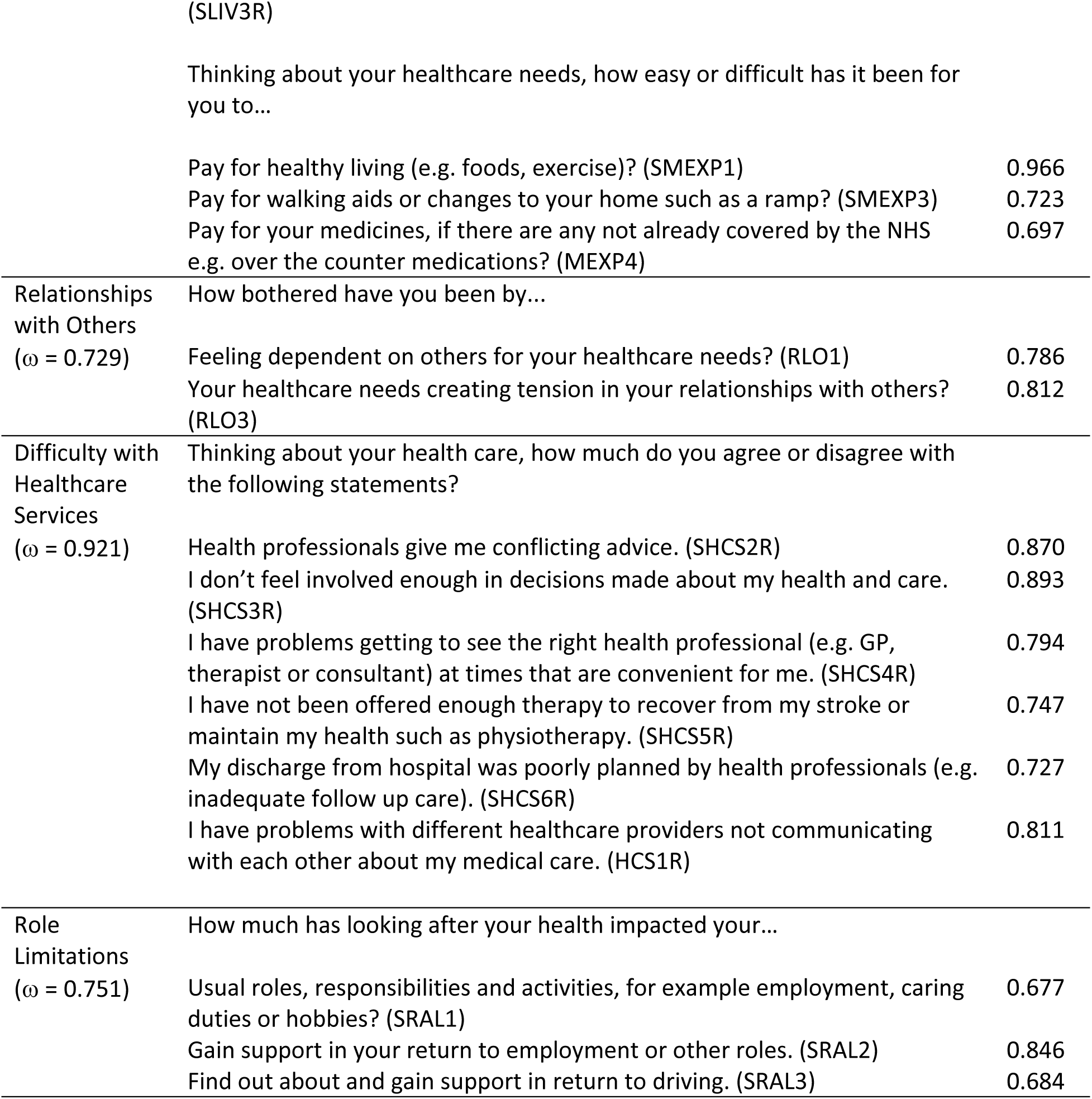
PETS-Stroke Factor Loadings and internal consistency for the 9 Factor (Best Fitting) Model.

### Test re-test reliability

Three hundred participants completed the PETS-stroke follow-up questionnaire. Test-retest reliability of the PETS-stroke was good for all domains (ICC>0.7). (**Table 4**)

### Construct validity

PETS-stroke domains were significantly correlated with measures of related constructs. Most domains of PETS-stroke demonstrated moderate correlations (Rho=0.4-0.59) with measures of stroke impact, and readiness to self-manage, and weak correlations (Rho=0.2-0.39) with satisfaction with care (**Table 4**). These correlations were consistent with our hypothesis that increased levels of treatment burden would be correlated with lower readiness to self-manage, lower satisfaction with stroke services and increased burden of illness, supporting the validity of the PETS-stroke.

Sensitivity analysis restricting analysis of domains ‘keeping healthy’ and ‘living at home with stroke’ to participants with full data only was fully consistent with our primary analysis (**Supplementary materials 4**).

**Table 4.** Correlations between PETS-stroke and other PRM’s and test-retest reliability of PETS-stroke.

| **PETS-stroke domain** | **SIS** | **SSMQ** | **PSQ** | **Test-retest ICC** |
| --- | --- | --- | --- | --- |
| Medical Information | -0.482** | -0.554** | -0.402** | 0.754 |
| Care Planning | -0.564** | -0.607** | -0.324** | 0.796 |
| Medications | -0.353** | -0.377** | -0.237** | 0.817 |
| Medical appointments | -0.559** | -0.463** | -0.302** | 0.799 |
| Keeping Healthy | -0.564** | -0.594** | -0.356** | 0.821 |
| Living at Home with Stroke | -0.428** | -0.503** | -0.330** | 0.835 |
| Relationships with Others | -0.586** | -0.453** | -0.226** | 0.776 |
| Difficulty with Healthcare services | -0.488** | -0.588** | -0.521** | 0.827 |
| Role Limits | -0.671** | -0.463** | -0.365** | 0.848 |
| Mental Fatigue | -0.604** | -0.452** | -0.248** | 0.807 |

Spearman's Rho; \*\*correlation significant at 0.01 level (two-tailed)
SSSQ: The Stroke Southampton Self-Management Questionnaire
PSQ: The Satisfaction with Stroke Care Measure
SIS: The Shortened Stroke Impact Scale
PRM: Patient Reported Measure
ICC: intra-class coefficient

### Known group validity

Higher treatment burden on PETS-stroke was significantly associated with poorer sociodemographic status in 8 out of 10 domains (**Table 5**). Participants from the most socioeconomically deprived areas had significantly higher treatment burden scores than participants from the least deprived areas, apart from in the ‘Medical information’ and ‘Role Limits’ domains. Participants who had 3 or more ongoing medical conditions had significantly higher treatment burden scores than those who had no co-morbidities in domains of Care Planning, Keeping Healthy, and Difficulty with Healthcare Services, but not in Mental Fatigue, Medical Information, Medications, Medical Appointments, Living at Home with Stroke, Relationships with Others, or Role Limits. (**Table 5**) Sensitivity analysis using Mann-Whitney U tests produced similar results. (**Supplementary materials 5**)

**Table 5.**
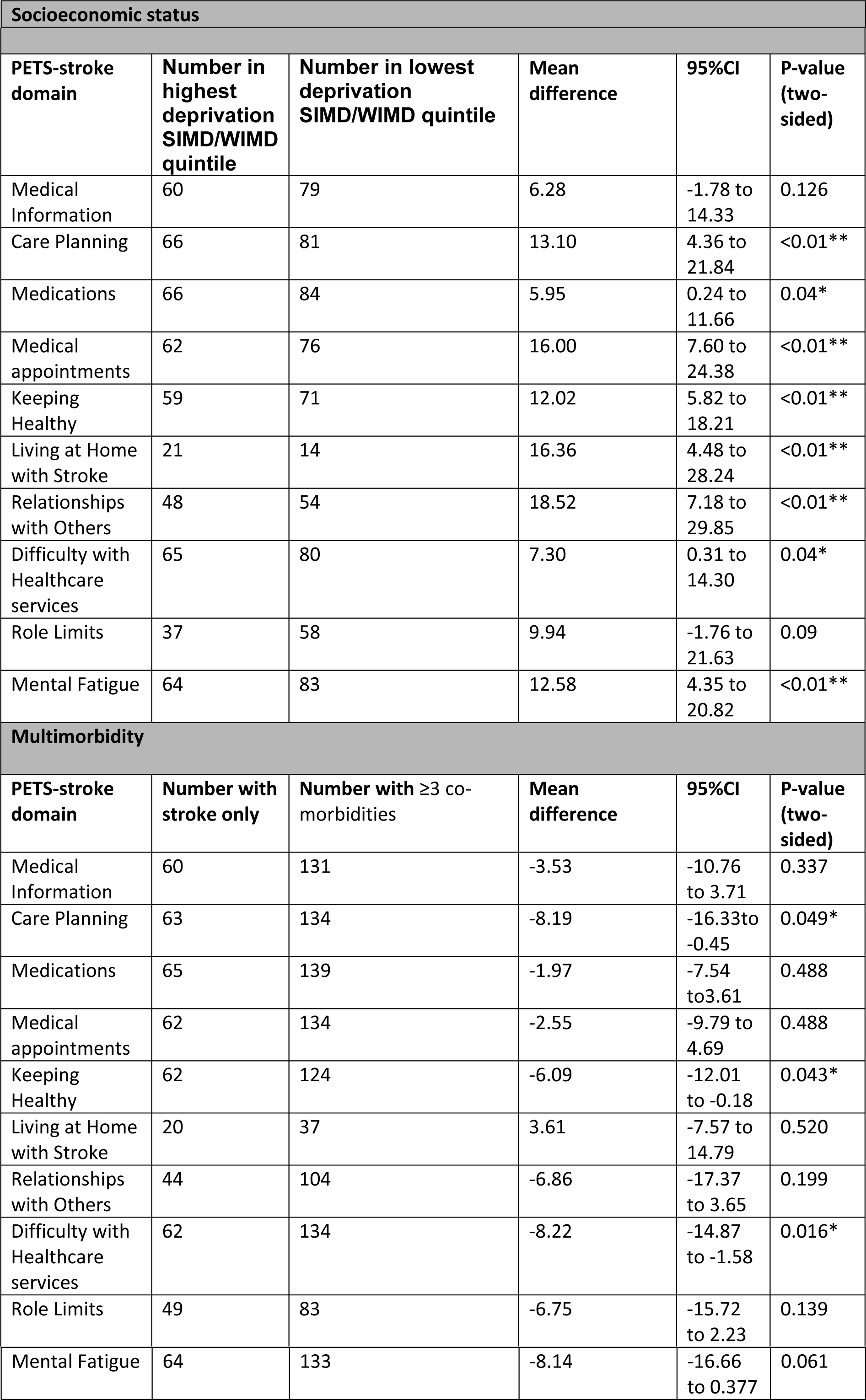
Difference in PETS-stroke domain scores based on socioeconomic and multimorbidity factors.

### Feasibility

Median reported duration to complete PETS-stroke was 15 (IQR=20) minutes, 108 participants (30%) reported engaging help to complete the questionnaire packs. A description of scores and missing data for each domain is shown in **Table 6**. For baseline assessment, number of missing items were within acceptable limits for 7/10 domains (**Table 6**). Living at home with stroke, Relationships with Others, and Role Limits had substantial missing items which prohibited scoring due to frequent selection of the ‘not applicable’ option. Possible floor effects were apparent for 5/10 domains (Medical Information, Care Planning, Medications, Medical Appointments, Difficulty with Healthcare Services). No domains demonstrated any ceiling effects. Feasibility data from follow-up assessments were consistent with baseline assessments and can be seen in **Supplementary materials 6**.

Thirty-one participants were interviewed. Participant demographics for the qualitative interviews can be found in **Supplementary material 7**.

Participants found the survey to be quick and easy to complete, with pen and pencil preferred [exemplar quotes: “…all quite self-explanatory. I didn’t have any problems”; “I would suggest the handwritten bit is better, especially for, eh, eh, well, a lot of people aren’t computer oriented”], though some suggested a digital option may be useful [exemplar quotes: “ sometimes I forget to look to my left, and then I’ve, I’ve realised there’s whole sections missing…..because I use my phone a lot, and I can make it big print…..and it can actually speak to me, if you get what I mean”; “would have happily filled it in online, I suppose that might have been easier to get back to you as well, if you know, if I couldn’t get to a post-box”]. The items contained within the questionnaire were important and relevant, but less relevant to those with mild stroke [exemplar quotes: “most of them were, yes…..I wouldn’t say none of them were, but I think the whole lot of them were kind of pretty relevant”; “I wasn’t required to have anything like that when….after the stroke….some of them more relevant than others”], and no important items were missing [exemplar quotes: “everything there, it compares, it compares to what I’ve, what I’m doing, what I’ve went through. And I’m finding the questionnaire, if the truth be told, I can’t think of anything else that I would really want to add to it”]. The questions were straightforward to understand but more difficult for those experiencing ongoing cognitive issues [exemplar quotes: “I’m very aware of the consequences of the stroke, so my sort of mental acuity, or cognitive ability has diminished radically…..if I didn’t understand the question, I could get (my partner) to explain it to me before I answered”]. Participants reported that the PETS-stroke provided them with a voice to express their opinions on their healthcare [exemplar quotes: “I don’t know about anybody else, but I felt as if there wasn’t a voice, if you get my meaning….”].

**Table 6.** Baseline PETS-stroke descriptives.

| **PETS-stroke domain** | **N baseline (total=381)** | **Mean score (SD)** | **Score Range** | **Missing (%)** | **% lowest score** | **% highest score** |
| --- | --- | --- | --- | --- | --- | --- |
| Medical Information | 351 | 30 (23) | 0-100 | 8% | 22.8% | 0.3% |
| Care Planning | 361 | 37 (27) | 0-100 | 5% | 19.4% | 2.8% |
| Medications | 373 | 21 (19) | 0-85 | 2% | 22.0% | 0.8% |
| Medical appointments | 353 | 31 (25) | 0-100 | 7% | 19.5% | 2% |
| Keeping Healthy | 329 | 35 (18) | 0-83 | 14% | 5.8% | 0% |
| Living at Home with Stroke | 89 | 47 (19) | 0-97 | 77% | 1.1% | 0% |
| Relationships with Others | 258 | 41 (31) | 0-100 | 32% | 15.9% | 7.8% |
| Difficulty with Healthcare Services | 363 | 35 (22) | 0-100 | 5% | 12.9% | 0.8% |
| Role Limits | 241 | 49 (28) | 0-100 | 37% | 8.3% | 6.2% |
| Mental Fatigue | 368 | 45 (26) | 0-100 | 3% | 13.0% | 4.6% |

### Changes to PETS-stroke and follow up content validity testing

Ten participants took part in cognitive interviews to discuss potential changes to the PETS-stroke. The potential changes discussed and results of analysis of the interviews are shown in **Supplementary materials 8 & 9**. Consensus was reached amongst authors that the resultant changes required were non-substantial, and that further testing was not required before recommendation of PETS-stroke as a valid tool.

## Discussion

### Summary and research in context

Our results support the validity of the PETS-stroke as a measure of treatment burden after stroke. Each domain of PETS-stroke demonstrated moderate to large correlations with at least one other self-report measure of stroke impact or care. The measure showed adequate test-retest reliability and internal consistency and was deemed acceptable and feasible to complete by stroke survivors. In the majority of domains, a higher level of treatment burden was reported in those who were more socioeconomically disadvantaged, similar to what has been demonstrated in other long-term conditions.^25^ However, in the majority of domains treatment burden was not significantly higher in stroke survivors with multimorbidity. While the literature on this association has been mixed,^26^ it is possible that these findings arose due to our low-fidelity method of measuring multimorbidity, necessary due to the limited range of multimorbidity within the study sample.

The factor structure that emerged for PETS-stroke differed to that observed in the PETS 2.0. In addition to 12 multi-item factors, the PETS 2.0 had 2 higher order factors reflecting healthcare workload and impact of workload;^16^ however, because of the refinement and adaptation of the PETS scale items to make them suitable for measuring treatment burden in stroke, it is unsurprising that the factor structure of this new scale differs from the original. This likely reflects the differing care pathway that stroke survivors experience relative to the multimorbid population. This emphasises the value of using a bespoke measure of treatment burden within the stroke population and suggests the PETS-stroke represents a distinct scale to the PETS 2.0, despite the PETS 2.0 serving as the basis for its design. Face validity testing had suggested that PETS-stroke is most relevant to those within one year of stroke, due to the emphasis on stroke rehabilitation. This work therefore tested the measure in that population. It is possible that measures developed for people with multimorbidity would be suitable for stroke survivors who experienced stroke a longer time ago if they are more focussed on the long-term management of their stroke alongside other conditions. Recently there have been several comprehensive patient-reported measures of treatment burden developed for use in multimorbidity,^27^ however PETS-stroke is the first stroke-specific measure.^28^ The PETS measure has been adapted for use in other contexts, for example for use in the homeless population^29^ and in kidney transplant patients.^30^ The development of a stroke-specific measure will allow inclusion of treatment burden as a case mix adjustor or outcome measure in clinical trials of stroke rehabilitation therapies. Improving the stroke patient experience was found to be a top ten priority in the recent James Lind Alliance priority seeking exercise in stroke.^31^ Previous research on patient experience in stroke has not tended to focus on treatment burden, rather on satisfaction with healthcare and unmet needs.^32^

### Strengths, limitations and future research

We have conducted a comprehensive evaluation of key psychometric properties of the PETS-stroke, surpassing the required sample size for all planned analyses. A key strength of our study is the range of disabilities and socioeconomic status reported by participants which will maximise the generalisability of the measure to the wider stroke population. Moreover, the content of this study aligns with the stroke survivor perspective, and involvement of primary care and stroke clinicians in the measure’s development enhances its potential for ‘real world’ use. PETS-stroke has been developed for use as a research tool, but future research could amend it for clinical use—this has been conducted with the original PETS measure.^33^

We did not assess responsiveness to change which is an important psychometric property necessary for assessing the impact of interventions, future work will address this.

The ethnic diversity of the sample population is limited. We therefore cannot say how well the PETS-stroke can be used by people whose first language is not English, or if there is any variability based upon cultural background.

Factor analysis revealed two items in the ‘living at home with stroke’ domain below optimal parameters, suggesting they could fit sub optimally in PETS-stroke. This domain also had a high proportion of participants ticking ‘not applicable’. Face validity testing had revealed these items to be irrelevant to some stroke survivors yet important to include in the measure.^6^ Future work should examine the sensitivity of the scale structure with and without those items.

Floor effects were apparent for half the items in the measure. This is likely a consequence of the study population being higher functioning than the typical stroke population. This may also explain some domains having a high number of ‘not applicable’ answered by participants. Similar floor effects have been seen with the development of other treatment burden measures.^34^

The variation in scale length for 2 of the PETS-stroke domains led to a minor skew of imputed scores, wherever data was missing. While our sensitivity analysis indicates that this skew did not have any meaningful impact upon our reported results, this issue should be addressed for future versions of PETS-stroke.

Our measure of multimorbidity was suboptimal and so findings of known groups validity testing in relation to that should be interpreted with caution. Further examination of treatment burden after stroke in relation to patient characteristics is important to identify those at high risk.

## Conclusions

In conclusion, our results indicate that the PETS-stroke is a valid measure of treatment burden in community dwelling stroke survivors. This new PRM has the potential to ascertain if treatments are workable for patients in the context of their everyday lives. Details of how the scale can be accessed and used can be found at http://tiny.cc/jgrlzz.

## Data Availability

Data is available upon reasonable request from the authors.

## Notes

### Competing Interest Statement

The authors have declared no competing interest.

### Clinical Trial

N/A

### Funding Statement

The project is funded by the Chief Scientist Office (CSO) Scotland (HIPS/21/13) and the Stroke Association (TSA LECT 2017_01). The funders had no role in study design, data collection and analysis, decision to publish, or preparation of the manuscript

### Author Declarations

Ethical approval has been provided on 16/9/20 by London and Surrey Borders NHS Research Ethics Committee (20/LO/0871).

